# A single centre randomised, feasibility study using point-of-care (POC) testing for respiratory viruses to direct oral corticosteroids use in preschool-aged children with acute wheeze. A Protocol

**DOI:** 10.1101/2025.08.25.25334354

**Authors:** Hannah Norman-Bruce, Clare Mills, Holly Drummond, Kathy Li, Hannah Mitchell, Lisa McFetridge, Mark Lyttle, Damian Roland, Ian Sinha, Thomas Waterfield, Helen Groves

**Author notes:** Name and contact information for the trial sponsor Kathryn Taylor, Research Governance Manager, Queen’s University Belfast, 63 University Road Belfast BT7 1NN, Northern Ireland. Role of sponsor/funders Queen’s University Belfast will act as Sponsor for the study and the Chief InvesUgator (CI) will take overall responsibility for the conduct of the study. Neither funding body (R&D department for HSCNI and *Asthma + Lung UK*) have a role in the study design, data acquisiUon, analysis and interpretaUon, or manuscript preparaUon. The views expressed are those of the author(s) and not necessarily those of the Funders.

## Abstract

**Background:** Wheezing in the pre-school aged group (under 5 years) is a common presentation and significant healthcare burden. It is a heterogenous presentation representing a spectrum of phenotypes and although the causes may be multifactorial, viral infection is the most common trigger, with rhinovirus and Respiratory Syncytial Virus (RSV) being the most commonly detected. Rigorous evidence-based guidance for the acute management of preschool wheeze (PW) with respect to which children likely to benefit from oral corticosteroid therapy (OCS), is lacking. RCTs of OCS use in PW have not adequately assessed the impact of respiratory virus testing in the management of PW. To address the hypothesis that OCS response may be determined by the specific virus, the feasibility of performing POC respiratory virus tests prior to randomisation in an acute paediatric ED setting needs to be ascertained.

**Methods:** The PRECISE Study will be a single centre, randomised, open-label, feasibility trial. Children aged 24-59 months with acute wheeze will be eligible if the clinician is uncertain if there is a role for oral corticosteroid therapy or not. At enrolment, participants will undergo a nasal swab for rapid respiratory virus testing. Children will be randomised in a 1:1 ratio to receive oral dexamethasone or not, based on their RSV result. Participants will continue to be managed by the clinician according to local guidance. They will be invited for clinical review at 72 hours where a repeat nasal swab may be performed. There will be a telephone follow up at one month and parents will be invited for extended telephone interviews within a further month. Comprehensive screening logs will address the primary outcome of recruitment and timeliness until enrolment. Remaining timeliness and adherence outcomes will be recorded in individual participant records and described using CONSORT diagrams. Acceptability will be measured using a mixed method qualitative approach based on the theory of acceptability framework.

**Discussion:** This pragmatically designed trial will address key feasibility points needed to inform a future, definitive multi-centre RCT prospectively testing the role of respiratory virus testing to randomise children with PW to receive oral corticosteroids or not.

**Trial registration:** NCT06580600 (clinicaltrials.gov)

## BACKGROUND AND RATIONALE

Wheezing in early childhood is very common, with over half of all children anticipated to have one episode of Preschool Wheeze (PW) by their sixth birthday (1,2) In the UK, cohort studies have consistently demonstrated a high prevalence of PW, with a recent increase in acute presentations. (3,4) Wheezing in the pre-school aged group is a heterogenous presentation representing a spectrum of phenotypes and multifactorial pathophysiology. (1,2) Viral infection is the most common trigger of acute PW. The most commonly implicated viruses are rhinovirus (RV) and Respiratory Syncytial Virus (RSV), each accounting for approximately a third of all infections. (5)

Oral corticosteroids are extensively used in the management of PW, despite little robust evidence to support their ubiquitous use. A recently published meta-analysis, including twelve PW trials conducted over the past two decades, failed to find a clinically meaningful benefit for OCS in PW. They were able to report a small reduction in wheeze severity at 4 hours (2.6%) but this was not sustained at 12 hours; they were also able to report a reduction, (approximately 3 hours) in length of stay for children receiving OCS.

**Table 1:**
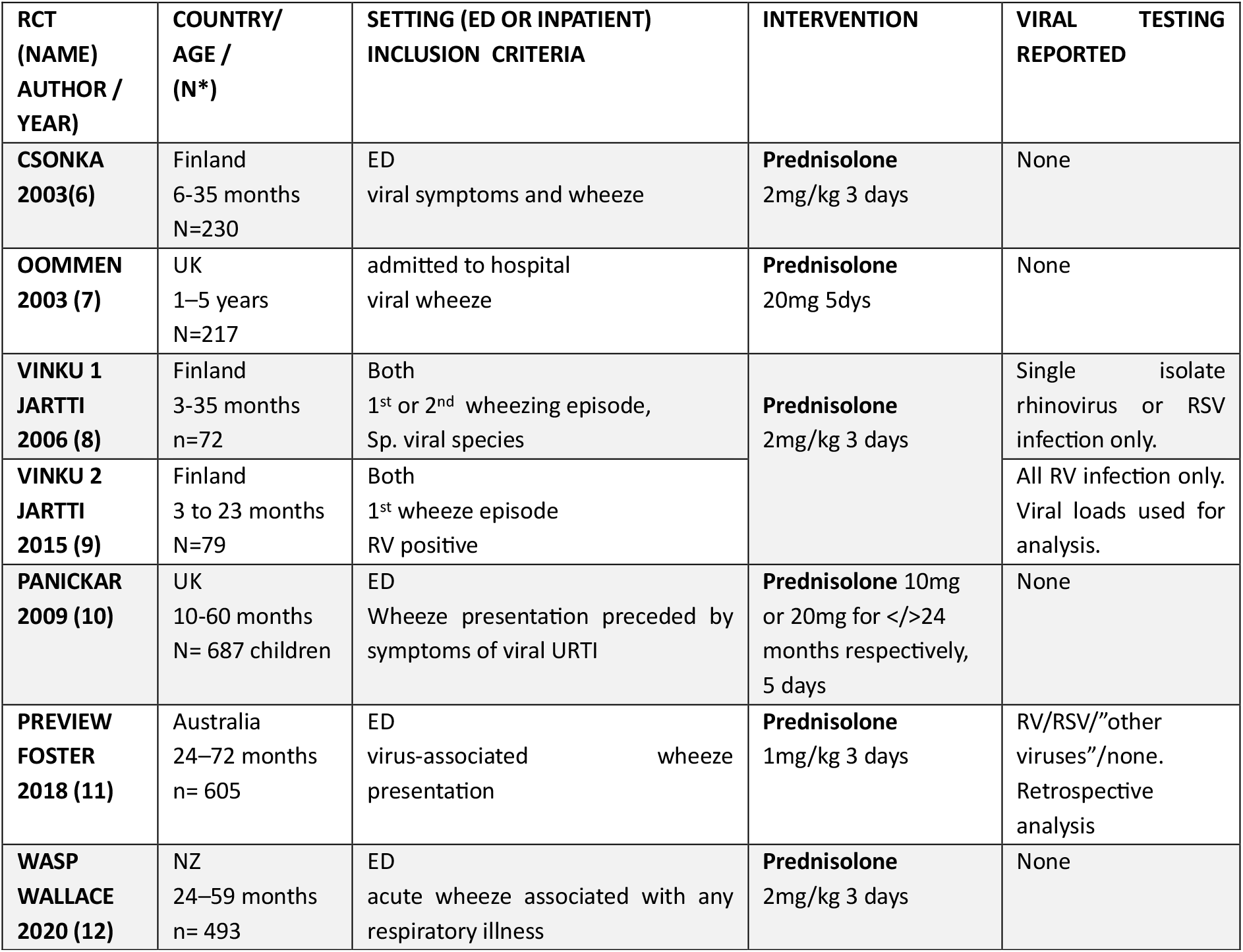
summary of RCTs for preschool wheeze since 2000, * Treated per protocol, RV = Rhinovirus, RSV Respiratory Syncytial virus, URTI = Upper respiratory tract infection.

One challenge of conducting clinical trials of OCS for children with PW is differentiating this from bronchiolitis. There is good evidence that OCS is not effective in the treatment of RSV-induced bronchiolitis, the commonest cause of wheezing and LRTI in under two-year-olds. (13,14) In previous studies of PW it has not been possible to prospectively exclude infants with proven RSV infection. This has led to many trials including infants with RSV infection, potentially masking the benefit of OCS for children with wheeze not due to RSV bronchiolitis.

Point-of-care respiratory viral testing is common in healthcare settings with results available within minutes. (15,16) However it remains unclear if POC respiratory viral testing can be used in a clinical trial setting to exclude children with RSV from a clinical trial of OCS for PW.

**We hypothesise that in children with mild-to-moderate acute PW, not associated with RSV, the use of OCS improves respiratory outcomes**

To address this hypothesis in a definitive trial, the acceptability and feasibility of performing POC respiratory pre-randomisation needs to be ascertained. The PRECISE Study is a single centre randomised trial to assess the feasibility of performing POC respiratory viral testing prior to OCS randomisation.

### AIMS AND OBJECTIVES

The overall aim is to inform the design of a future, definitive, multicentre RCT comparing OCS to no OCS for acute PW, using POC respiratory viral testing to selectively identify and cohort eligible participants.

#### Primary objective

- To evaluate the feasibility of randomisation to receive OCS or no OCS treatment based on POC viral testing results in pre-school aged children with wheeze in a paediatric ED setting

#### Secondary objectives

- To evaluate barriers to enrolment of patients in a future, definitive trial
- To describe viral aetiology of preschool wheeze in included cohort
- To assess parent/guardian acceptability of using a viral POC test in ED to formulate a management plan for PW
- To assess the feasibility of obtaining a second follow up nasal swab (NS) and achieving follow up at specified time points

#### Exploratory objectives

- To assess the feasibility of obtaining finger prick blood test for POC testing of peripheral blood eosinophil counts in a paediatric ED setting
- Describe peripheral eosinophil count values in patients undergoing optional testing

### TRIAL DESIGN & SETTING

This feasibility study will be conducted as a single-centre, prospective, randomised open-label study. The flow diagram depicting an overview of the study, including the optional components of the study, is presented in Figure 1. The study site will be solely the Royal Belfast Children’s Hospital, (RBHSC). Recruitment will take place in the RBHSC Emergency Department (ED).

**Figure 1.**
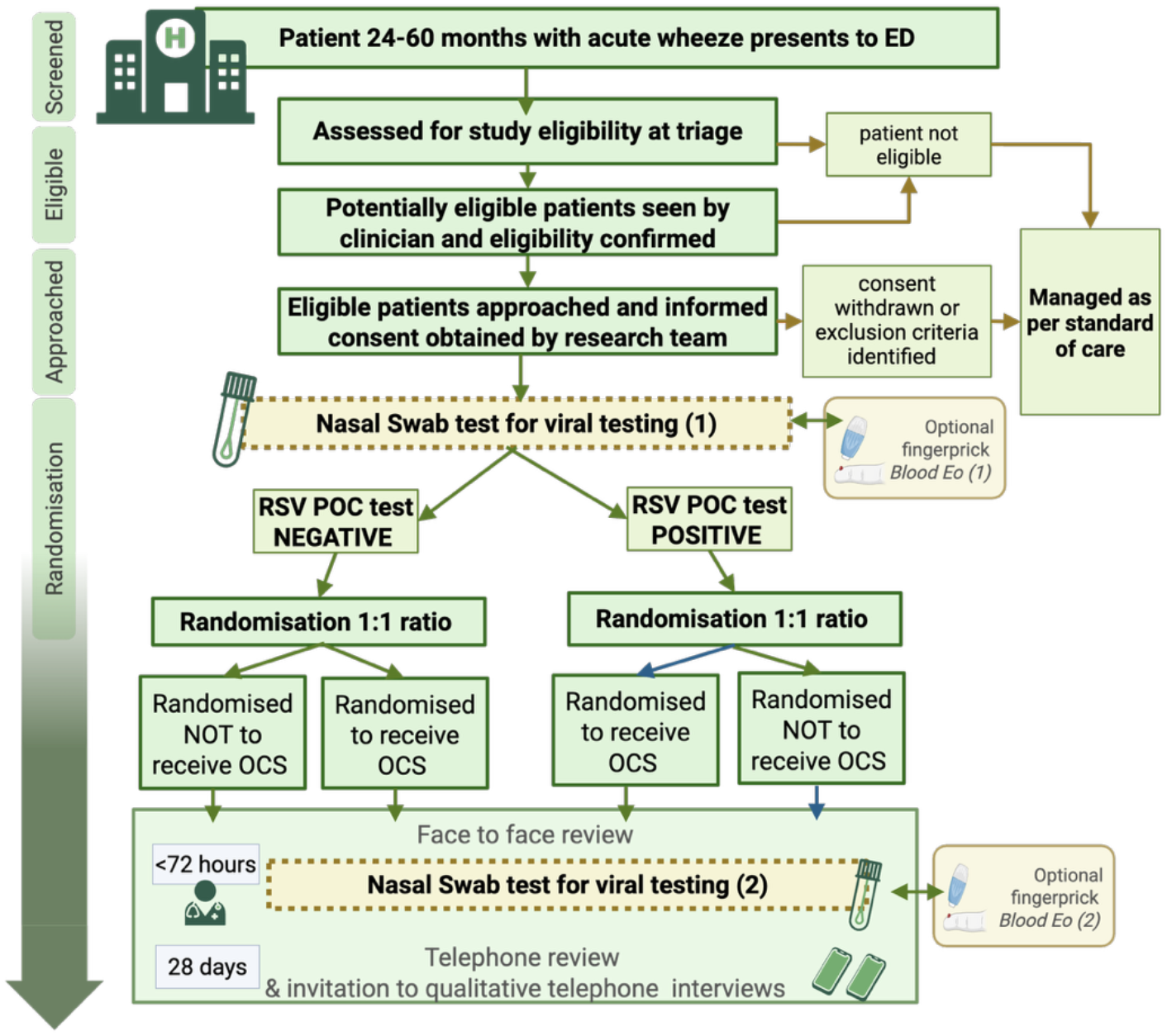
summary of flow of participants through the PRECISE Study. Created with BioRender.com

### POPULATION / ELIGIBILITY CRITERIA

Participants meeting study eligibility criteria by age and clinical diagnosis will be entered anonymously into a screening log in the ED. Eligibility will be confirmed by a member of the clinical team using the eligibility criteria and subsequently confirmed by a member of the research team who is named on the delegation log.

### Inclusion Criteria

‐ Aged 24-60 months
‐ Clinical diagnosis of acute wheeze

### Exclusion Criteria

‐ OCS definitely required following clinician assessment
‐ OCS definitely not required following clinician assessment
‐ Signs and symptoms of severe or life-threatening wheeze (Table 2)
‐ Patients presenting with wheeze suspicious for a non–respiratory cause
‐ Clinical evidence of shock (e.g. prolonged capillary refill time greater than 3 seconds)
‐ Clinical evidence of bacterial sepsis as indicated by the RBHSC sepsis screening tool
‐ Past history of severe or life-threatening asthma or history of previous intensive care admission with acute wheeze
‐ History of preterm birth (before 30 weeks gestation) or with a diagnosis of chronic lung disease
‐ Known immunodeficiency/ongoing immunosuppressive therapy
‐ Contraindication to oral corticosteroids
‐ Previously enrolled in the PRECISE Study
‐ Child not assessed by paediatric clinician acting as decision maker on the middle grade rota
‐ Refused POC nasal swab testing

**Table 2:**
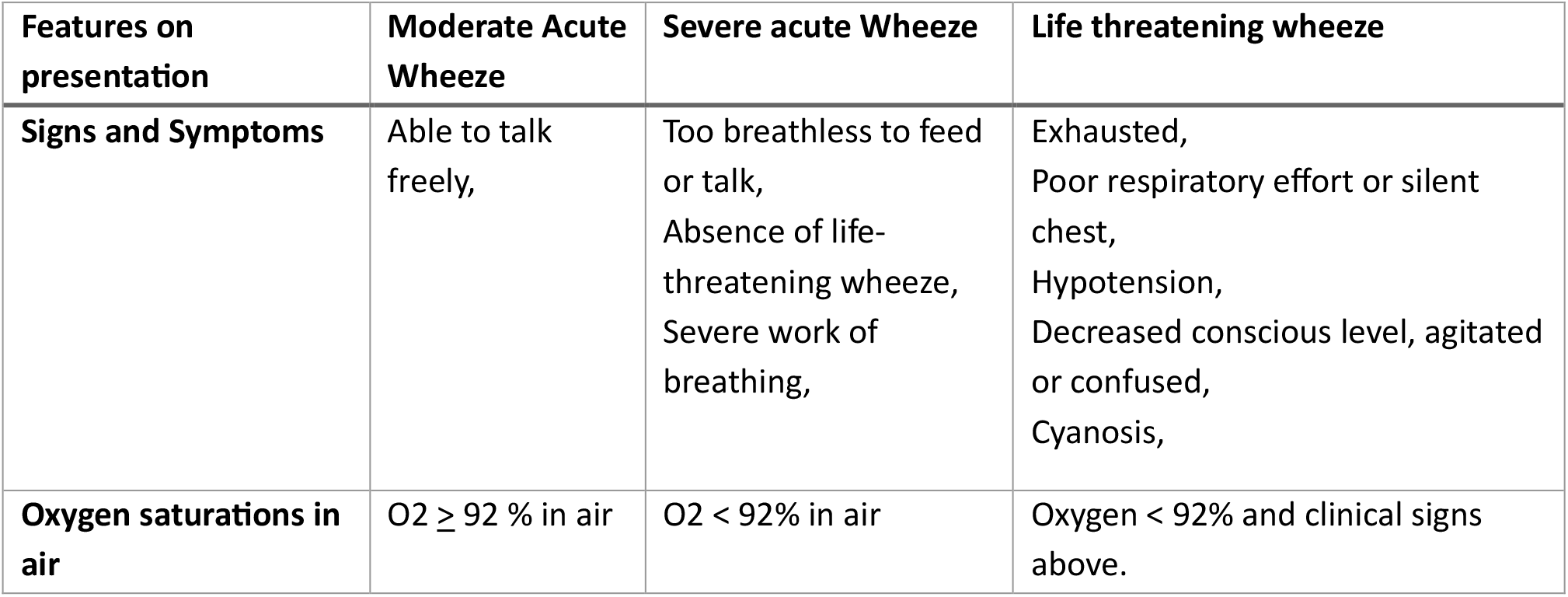
Summary of wheeze severity (Adapted from BTS/SIGN guidance)(17)

### CONSENT AND ENROLMENT

Potential participants will be identified by clinicians and invited to speak with a member of the research team. The research team will then approach the parent/guardian(s) of potentially eligible patients, confirm eligibility, provide written participant information and discuss the trial. This includes detailing the optional components of the study and potential to use nasal swab samples in ancillary studies. A screening log of all patients meeting the inclusion criteria will be maintained and constructed based upon the SEAR framework.(18)

The parent/guardian(s) of potentially eligible patients will be allowed a maximum of 60 minutes to decide if they wish their child to be included in the trial. Written consent will be taken by an authorised member of the training log, from a legal parent/guardian(s) on behalf of the participant. The consent form clearly denotes the use of swabs. A log of those declining consent, including the reasons for this, will be maintained.

### STUDY PROCEDURES

#### Nasal swab event

Sampling for respiratory virus testing will occur at initial presentation and at the two-day follow-up. Nasal swabs will be performed by the research team in a standardised method using a minitip flocked swab (COPAN ITALIA SpA) in each nostril. The material will initially be tested using the cobas^®^ liat system [Roche Diagnostics] which uses multiplex RT-PCR assay for rapid detection of RSV and influenza A/B. Residual material will be sent to the Regional Virus laboratory for identification of any additional viral pathogens (SARS-CoV-2, seasonal coronaviruses, rhinovirus, adenovirus, human metapneumovirus and parainfluenza viruses). Any remaining residual nasal swab material will be frozen and stored for further analysis of airway immune responses at Queen’s University Belfast (QUB). All samples transferred to QUB will be anonymised prior to transfer. Nasal swabbing is commonly performed for children being admitted to hospital and will be performed by experienced practitioners to minimise any discomfort/distress for the participant and their parent/guardian(s).

#### Finger prick eosinophil count

For participants who consent to optional finger prick blood testing for peripheral blood eosinophils (PBE), this will be performed at the same time as the nasal swab. PBE counts will be obtained using POC testing on the Hemocue® WBC DIFF device [Accuscience] which provides a 5-point full white cell differential result.

Results of the POC tests, extended viral testing, and peripheral blood eosinophils will be made available to the family and their clinical team, who are familiar with interpreting and actioning such results, and documented by the research team in the child’s medical record.

### RANDOMISATION AND ALLOCATION

Participants will be randomised using an automated web-based system using permuted blocks in a 1:1 ratio according to RSV test results. Randomisation will be completed on the REDCap platform, used for the completion of electronic Case Report Forms (CRF) by an appropriately trained and delegated member of the research team. The allocation sequence is designed by the trial co-ordinator and will not be shared with wider teams to avoid prediction of randomisation and selective enrolment.

### BLINDING

Parents/guardians, those who provide health care to the participants, and outcome assessors, will *not* be blinded to the allocated intervention. This includes the study statistician, who has no role in decision-making with regards the conduct of the study, and the remainder of the research team will also be unblinded for the purposes of managing data collection and reviewing cases to assess for protocol deviations.

### INTERVENTION

Oral corticosteroid is usually given in the RBHSC ED as oral dexamethasone (typically 0.3mg/kg). It is given as a single dose therefore adherence is not an issue in the acute setting. If the participant vomits within 30 minutes of dexamethasone administration, it may be re-administered at the discretion of the treating clinician and recorded in the CRF. The unblinded clinician may choose to determine the OCS therapy plan at any point after enrolment, irrespective of allocation (e.g prescribe OCS to a child randomised not to receive OCS). Any deviation from planned study randomisation will be recorded on the CRF. After randomisation participants will continue to follow this RBHSC guideline, there will be no further difference in-between arms. Clinicians may perform investigations and administer additional treatment according to local hospital guidance. These, and any other concomitant medications, will be recorded on the CRF.

### OUTCOME MEASURES AND CRITERION FOR ASSESSING FEASIBILITY OF TRIAL

*The Primary outcomes are listed in the table 3 alongside thresholds of feasibility for this study design to inform a future randomized controlled trial. Thresholds were determined by relevant outcomes used in randomised controlled trials for the management of preschool wheeze and existing literature for the design of feasibility studies.* (19–21) *Thresholds were incorporated into traffic light criteria with SMG and expert PPI consensus. Where appropriate, thresholds are defined as proportions or percentages of participants so that all four outcomes can be evaluated together. In order to progress to a full trial without substantial changes in design, the trial must meet all the pre-specified quantitative thresholds with a 10% margin.*

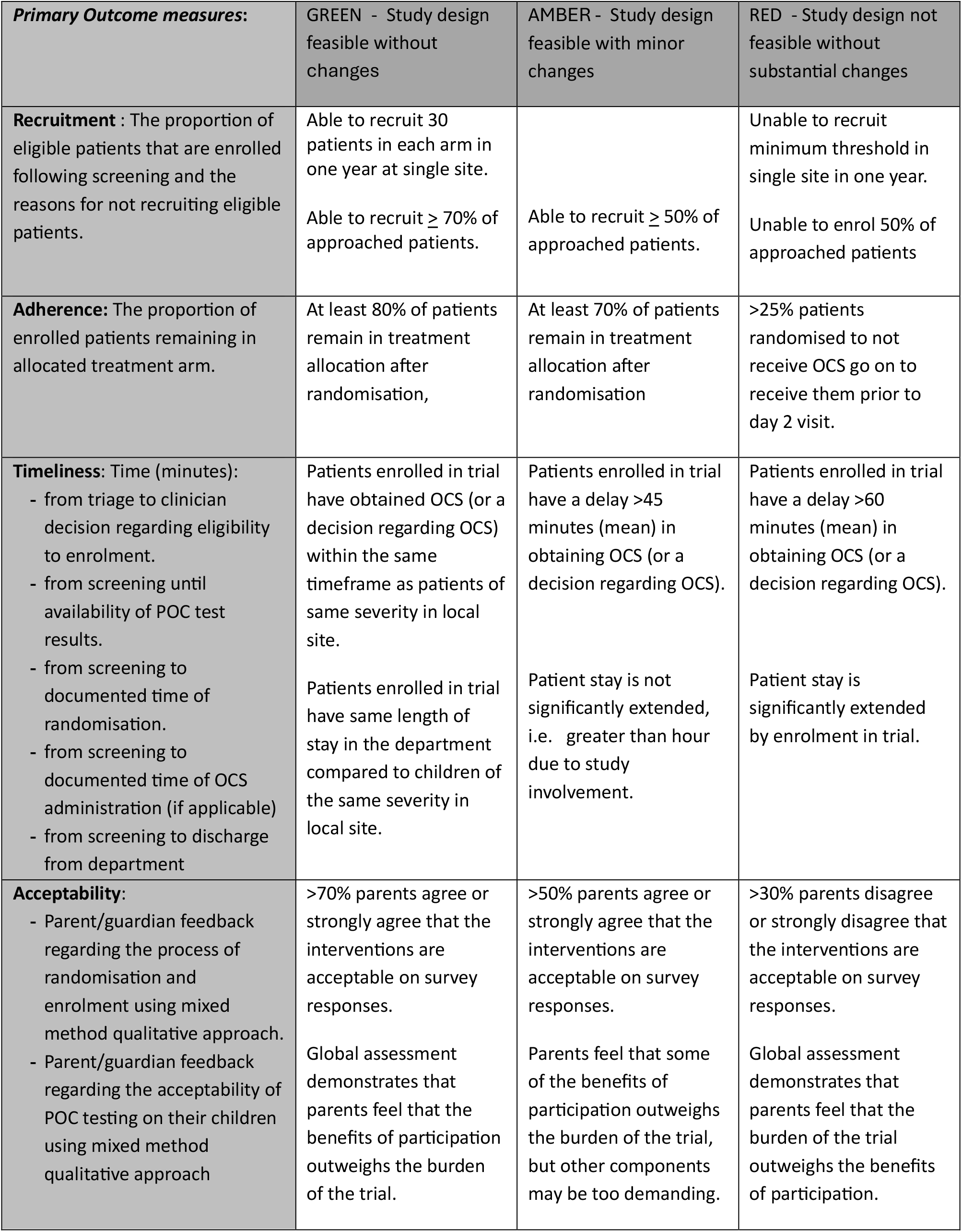

| **Primary Outcome measures:** | GREEN - Study design feasible without changes | AMBER - Study design feasible with minor changes | RED - Study design not feasible without substantial changes |
| --- | --- | --- | --- |
| **Recruitment** : The proportion of eligible patients that are enrolled following screening and the reasons for not recruiting eligible patients. | Able to recruit 30 patients in each arm in one year at single site.<br>Able to recruit $\geq 70\%$ of approached patients. | Able to recruit $\geq 50\%$ of approached patients. | Unable to recruit minimum threshold in single site in one year.<br>Unable to enrol 50% of approached patients |
| **Adherence:** The proportion of enrolled patients remaining in allocated treatment arm. | At least 80% of patients remain in treatment allocation after randomisation, | At least 70% of patients remain in treatment allocation after randomisation | >25% patients randomised to not receive OCS go on to receive them prior to day 2 visit. |
| **Timeliness:** Time (minutes):<br>- from triage to clinician decision regarding eligibility to enrolment.<br>- from screening until availability of POC test results.<br>- from screening to documented time of randomisation.<br>- from screening to documented time of OCS administration (if applicable)<br>- from screening to discharge from department | Patients enrolled in trial have obtained OCS (or a decision regarding OCS) within the same timeframe as patients of same severity in local site.<br>Patients enrolled in trial have same length of stay in the department compared to children of the same severity in local site. | Patients enrolled in trial have a delay >45 minutes (mean) in obtaining OCS (or a decision regarding OCS).<br>Patient stay is not significantly extended, i.e. greater than hour due to study involvement. | Patients enrolled in trial have a delay >60 minutes (mean) in obtaining OCS (or a decision regarding OCS).<br>Patient stay is significantly extended by enrolment in trial. |
| **Acceptability:**<br>- Parent/guardian feedback regarding the process of randomisation and enrolment using mixed method qualitative approach.<br>- Parent/guardian feedback regarding the acceptability of POC testing on their children using mixed method qualitative approach | >70% parents agree or strongly agree that the interventions are acceptable on survey responses.<br>Global assessment demonstrates that parents feel that the benefits of participation outweighs the burden of the trial. | >50% parents agree or strongly agree that the interventions are acceptable on survey responses.<br>Parents feel that some of the benefits of participation outweighs the burden of the trial, but other components may be too demanding. | >30% parents disagree or strongly disagree that the interventions are acceptable on survey responses.<br>Global assessment demonstrates that parents feel that the burden of the trial outweighs the benefits of participation. |

**Table 3:**
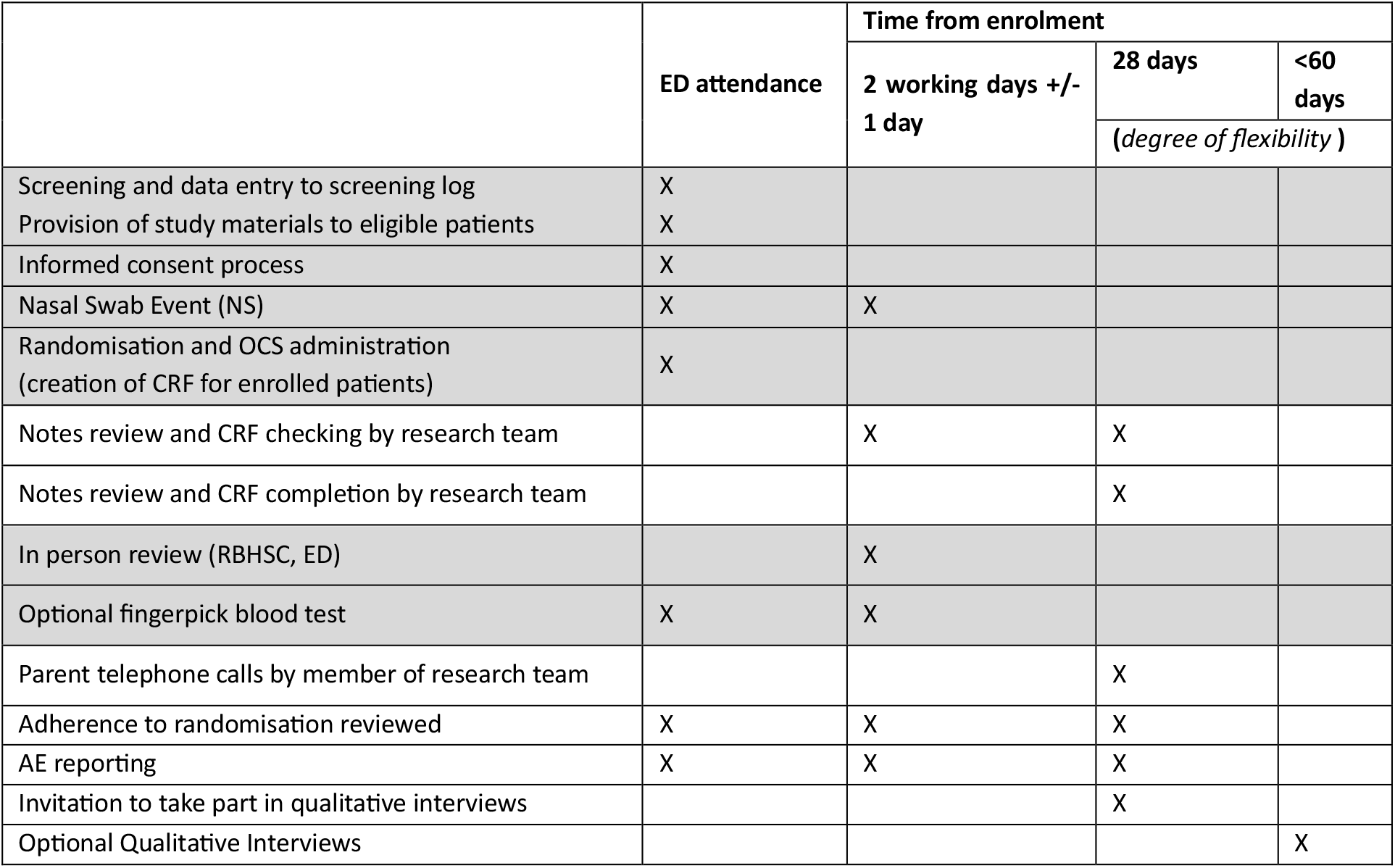
Schedule of assessments (grey, denotes in person activities, white, denotes remote activities)

|  | ED attendance | Time from enrolment |  |  |
| --- | --- | --- | --- | --- |
|  |  | 2 working days +/- 1 day | 28 days | <60 days |
|  |  |  | (degree of flexibility ) |  |
| Screening and data entry to screening log | X |  |  |  |
| Provision of study materials to eligible patients | X |  |  |  |
| Informed consent process | X |  |  |  |
| Nasal Swab Event (NS) | X | X |  |  |
| Randomisation and OCS administration (creation of CRF for enrolled patients) | X |  |  |  |
| Notes review and CRF checking by research team |  | X | X |  |
| Notes review and CRF completion by research team |  |  | X |  |
| In person review (RBHSC, ED) |  | X |  |  |
| Optional fingerpick blood test | X | X |  |  |
| Parent telephone calls by member of research team |  |  | X |  |
| Adherence to randomisation reviewed | X | X | X |  |
| AE reporting | X | X | X |  |
| Invitation to take part in qualitative interviews |  |  | X |  |
| Optional Qualitative Interviews |  |  |  | X |

#### Secondary outcome measures

‐ Proportions of patients with positive respiratory viral testing results for RSV and/or other respiratory viruses, or no virus detected
‐ Proportion of discordant viral swab results between consecutive tests in the same patient
‐ Proportion of enrolled patients completing second nasal/NP swab, follow up assessments and completeness of recording of follow-up information

#### Exploratory outcome measures

‐ Proportion of enrolled patients completing optional finger prick blood testing
‐ Description of mean peripheral eosinophil count values in enrolled patients

### DATA COLLECTION & ASSESSMENT SCHEDULE

Overall, the study incorporates three follow up points per participant as summarised in the schedule of assessments (Table 3). UK BTS/SIGN guidance recommends all patients presenting with acute asthma or wheeze should be reviewed following discharge from ED/hospital within two working days.(17) Participants will be offered an in person review with a clinical member of the research team within two working days (+/−1 day) in the ED department. This review will include a brief, standardised clinical assessment and clinical data will be collected in line with standard care for ED review visits.

The parent/guardian(s) will be telephoned by a member of the research team approximately 28 days after enrolment to complete a survey about participant outcomes as well as adherence and initial review of trial acceptability. This review will be pre-scheduled with the parent/guardian(s) at the day two appointment. If a participant does not present for planned in person review, the research team will aim to contact parent/guardian(s) up to three separate times by telephone on the same day. If unsuccessful, they will try the following day or using an alternative phone number if provided. If still unsuccessful, an email invitation to contact the research team will be sent. If no further contact is achieved, the participant will be marked as lost to follow up. Remuneration for the burden of time caused by the PRECISE Study will be provided to the families through the use of an online gift voucher sent by email.

All data will be collected and recorded in electronic case report forms (CRF) within a bespoke, web-based, electronic platform using REDCap software. (22) Hard copies of consent forms will be used, and kept securely alongside the participant linkage log. The screening logs and electronic CRFs will capture data pertaining to the primary outcomes. The screening log will capture the recruitment rates and the timeliness of patient flow through the trial up to recruitment and randomisation. The remaining timeliness data will be captured in individual CRFs alongside adherence over the three time points in the study period.

Acceptability of the trial interventions will be measured using a ten-point questionnaire based on the Theory of Acceptability framework.(23) This will be sent electronically to parents/guardians on day 2 and day 28 review, but it can be read out to people without access to emails or difficulties reading. Additionally, parent/guardian(s) of enrolled patients will be invited to participate in exploratory qualitative interviews within two months of enrolment.

### DATA MANAGEMENT & CONFIDENTIALITY

Participant data will be held within the secure REDCap platform linked to Queen’s University Belfast. The research team and site staff will undergo training to ensure all data is accurate, complete, and reliable. Quality is also assured through the use REDCap software allowing full audit trail, consistency, and compliance with GCP and relevant regulatory requirements.(22) For routinely collected clinical data from clinical records, researchers will use the participant study number (PSN) assigned at randomisation to link data pseudo - anonymously. Regular data review will be carried out by the study co-ordinator and PI to ensure consistency and identify any discrepancies or protocol deviations. Additionally, quality control will be ensured by the use of Standard Operating Procedures (SOPs) of the Belfast Trust. Data will only be accessed by delegated members of the research team, a list of which will be kept in the Study Master File (SMF). All essential documents including participant consents and linkage logs will be stored in a study file in locked cabinets according to Belfast Trust data storage and archiving guidelines.

### SAMPLE SIZE & RECRUITMENT

The PRECISE study will recruit a minimum of 30 participants testing positive for RSV and 30 participants testing negative for RSV. However, as RSV status will be determined after enrolment the PRECISE Study has capacity to enrol up to 150 participants to ensure minimum requirements are met.

A target of 30 participants per group is in line with literature for feasibility studies using mixed methodology. (19,21,24) In order to truly address the primary outcome of acceptability in this feasibility study it is imperative that the voice of families is represented. A minimum of 60 participants was informed by an anticipated attrition rate to complete surveys (50%) and uptake of qualitive interviews (15%) from previous studies. (25) This needs to be achieved in both treatment arms to ensure that adherence is not different depending on viral status. Feasibility of achieving this target is supported by local audit data of ED attendances in previous winter seasons.

### DATA ANALYSIS

The progress of all eligible participants will be described, from screening to completion of the study, using a CONSORT flow diagram. Descriptive statistics for continuous variables (mean, inter quartile ranges) and categorical variables (frequency counts, percentages) will be used to summarise baseline characteristics of screened and enrolled patients. Potential barriers to recruitment will be investigated by comparing the descriptive statistics between those recruited and those eligible but not recruited using chi-squared or Fisher’s exact tests for categorical data and Mann-Whitney U tests for continuous data. The proportion of participants who successfully complete consent, randomisation, receipt of intended treatment allocation, and evaluation for each objective, will be described as percentages of the total number screened for eligibility and approached. The reasons given for not recruiting eligible patients will be collated and described. The proportion of enrolled patients remaining in their allocated treatment arm will be described as percentages of the total number randomised. The time taken for each stage of patient flow will be described as medians (with interquartile ranges) and graphically using Kaplan-Meier plots.

Quantitative secondary outcome analysis will also be descriptive. The proportion of patients undergoing second NS and finger prick blood tests will be reported, and the viral pathogens detected on all point-of-care testing will be described along with CT values of positive tests. The proportion of discordant viral swab results between consecutive tests in the same patient will be reported. The study has been designed to minimise missing data for the primary analysis; missing data will be excluded from the analyses.

Qualitative interview data will be transcribed internally, checked, and anonymised as the study progresses. QSR NVivo software will be used to assist in the organisation and indexing of qualitative data. Data will be analysed thematically, and analysis will be informed by the constant comparison approach of grounded theory. (26) Both the interview topic guides, and the parental surveys have been adapted from the framework of acceptability to facilitate a robust mixed methods analysis to address acceptability outcomes. (23) Parent/guardian(s) will complete surveys prior to the additional interviews and their responses will be available to the interviewer. This is designed for triangulation of survey responses; however, the interviews will be able to support development of new ideas, particularly regarding ways to improve the study design, and may identify themes beyond the framework which may not have been identified by the researchers.

### SAFETY, ETHICS AND GOVERNANCE

Recording adverse events associated with OCS use such as rash/allergic reaction, vomiting, or behaviour change, represents standard care. Any adverse events reported as part of routine clinical care will be summarised and reported.

The PRECISE Study protocol [Protocol V3 18/11/2024] has received a favourable opinion from the Health and Social Care Research Ethics Committee A [Reference 24/NI/0050]. The investigators will conduct the study in compliance with the protocol that was given approval/favourable opinion by the REC. Any protocol deviations will be documented and escalated as appropriate. Of note, the open-label nature of this study is such that final decision for delivery (or not) of OCS is at the discretion of the treating clinician. Indeed, adherence to treatment arm is an outcome measure of feasibility, and non-adherence to treatment arm is not considered a breach of protocol compliance. Patient confidentiality and data protection is held with utmost importance and as described, data management and sample handling will comply with necessary regulations.

The Study Management Group (SMG) is comprised of PI, study co-ordinator, an independent advisor from the Northern Ireland Clinical Trials Unit (CTU), and three other co-investigators who provide study specific expertise. The SMG meet at least quarterly by teleconference and will communicate between times via email as needed. Meetings will be formally minuted and stored in the SMF. As described above, regular data review will be carried by the study co-ordinator and available to audit by the Belfast Trust and sponsor.

### DISSEMINATION

The findings of the PRECISE study will be reported in accordance with the CONSORT guidelines and will aim to be published in high quality, externally peer-reviewed, open-access journals. (27) The study findings will be presented at national and international meetings and disseminated through established connections with paediatric emergency medicine and respiratory medicine partners. Furthermore, the study co-ordinator and PI will engage with Asthma + Lung UK and the Study’s PPI Advisory Group to produce lay summaries and determine a strategy to disseminate findings to the public. This will secure broad stakeholder engagement with the design for the future definitive trial and allow the results will be readily accessible to all stakeholders.

Authorship, for each publication and presentation, will be determined according to the internationally agreed criteria for authorship. The study will comply with the good practice principles for sharing individual participant data from publicly funded research and data sharing will be undertaken in accordance with the required regulatory requirements(28).

## DISCUSSION

Evaluating the role of POC respiratory virus testing to predict OCS responsiveness in the acute management of PW is a research priority. In order to test the hypothesis that in a subgroup of children with acute PW, not associated with RSV, use of OCS will improve respiratory outcomes, feasibility of such intervention in an acute ED setting needs to be ascertained.

The PRECISE study will evaluate barriers to enrolment of patients in a future, definitive trial. The study will evaluate the feasibility and acceptability of using POC viral swab results in order to randomise children to receive OCS or not. Additionally, to our knowledge, this is the first study to evaluate the role of POC testing for peripheral blood eosinophils in the acute paediatric ED setting for preschool population. The PRECISE study will report the viral aetiology and peripheral blood eosinophil counts of preschool aged children with wheeze enrolled in the trial.

## Data Availability

Study materials are available upon reasonable request. Study data will be made publicly available upon final publication in university repository.

## Author Contributions & Competing interests

HG, HNB and TW were involved in trial conceptualisation, design and execution. CM, HD and KL provided expertise and provide support processing viral tests and transferring to QUB. DR, ML and IS are SMG members who have supported design and provided specialist clinical expertise. HL and LMcF have provided advice on statistical analysis. All authors have reviewed final manuscript. All authors and those mentioned in acknowledgements have given consent for publication. None of the study investigators or authors have any competing interests to declare.

## Ethics approval

The PRECISE Study has received a favorable opinion from the Health and Social Care Research Ethics Committee A [Reference 24/NI/0050].

## Acknowledgements

We would like to thank the PPI co-lead, Mrs E. Houldsworth, and the PERUKI network for their invaluable support with the PRECISE study.

## TRIAL STATUS

Precise Study has been recruiting patient in the RBHSC since 4/11/2024.

## Abbreviations

BHSCT: Belfast Health and Social Care Trust
CONSORT: Consolidated Standards of Reporting Trials
CRF: Case Report Form
ED: Emergency Department
GCP: Good Clinical Practice
NS: Nasal Swab
NHS: R&D National Health Service Research & Development
OCS: Oral corticosteroid (Therapy)
PI: Principal Investigator
POC: Point-of-care
PW: Pre-school Wheeze
PIS: Participant Information Sheet
PPI: Patient and Public involvement
RCT: Randomised Control Trial
REC: Research Ethics Committee
RV: Rhinovirus
RBHSC: Royal Belfast Hospital for Sick Children
RSV: Respiratory syncytial virus
SOP: Standard Operating Procedure
SMF: Study Master File
SMG: Study Management Group

## References

1. Brand PLP, Baraldi E, Bisgaard H, Boner AL, Castro-Rodriguez JA, Custovic A, et al. Definition, assessment and treatment of wheezing disorders in preschool children: an evidence-based approach. European Respiratory Journal. 2008 May 14;32(4):1096–110.

2. Martinez FD, Wright AL, Taussig LM, Holberg CJ, Halonen M, Morgan WJ. Asthma and Wheezing in the First Six Years of Life. N Engl J Med. 1995 Jan 19;332(3):133–8.

3. Bloom CI, Franklin C, Bush A, Saglani S, Quint JK. Burden of preschool wheeze and progression to asthma in the UK: Population-based cohort 2007 to 2017. Journal of Allergy and Clinical Immunology. 2021 May;147(5):1949–58.

4. Belgrave DCM, Granell R, Turner SW, Curtin JA, Buchan IE, L. Souëf PN, et al. Lung function trajectories from pre-school age to adulthood and their associations with early life factors: a retrospective analysis of three population-based birth cohort studies. The Lancet Respiratory Medicine. 2018 July;6(7):526–34.

5. Kengne–Nde C, Kenmoe S, Modiyinji AF, Njouom R. Prevalence of respiratory viruses using polymerase chain reaction in children with wheezing, a systematic review and meta–analysis. Shields MD, editor. PLoS ONE. 2020 Dec 14;15(12):e0243735.

6. Csonka P, Kaila M, Laippala P, Iso-Mustajärvi M, Vesikari T, Ashorn P. Oral prednisolone in the acute management of children age 6 to 35 months with viral respiratory infection-induced lower airway disease: a randomized, placebo-controlled trial. The Journal of Pediatrics. 2003 Dec;143(6):725–30.

7. Oommen A, Lambert PC, Grigg J. Efficacy of a short course of parent-initiated oral prednisolone for viral wheeze in children aged 1–5 years: randomised controlled trial. The Lancet. 2003 Nov;362(9394):1433–8.

8. Jartti T, Lehtinen P, Vanto T, Hartiala J, Vuorinen T, Mäkelä MJ, et al. Evaluation of the efficacy of prednisolone in early wheezing induced by rhinovirus or respiratory syncytial virus. Pediatr Infect Dis J. 2006 June;25(6):482–8.

9. Jartti T, Nieminen R, Vuorinen T, Lehtinen P, Vahlberg T, Gern J, et al. Short- and long-term efficacy of prednisolone for first acute rhinovirus-induced wheezing episode. J Allergy Clin Immunol. 2015 Mar;135(3):691–698.e9.

10. Panickar J, Lakhanpaul M, Lambert PC, Kenia P, Stephenson T, Smyth A, et al. Oral Prednisolone for Preschool Children with Acute Virus-Induced Wheezing. N Engl J Med. 2009 Jan 22;360(4):329–38.

11. Foster SJ, Cooper MN, Oosterhof S, Borland ML. Oral prednisolone in preschool children with virus-associated wheeze: a prospective, randomised, double-blind, placebo-controlled trial. The Lancet Respiratory Medicine. 2018 Feb;6(2):97–106.

12. Wallace A, Sinclair O, Shepherd M, Neutze J, Trenholme A, Tan E, et al. Impact of oral corticosteroids on respiratory outcomes in acute preschool wheeze: a randomised clinical trial. Arch Dis Child. 2021 Apr;106(4):339–44.

13. Ricci V, Nunes VD, Murphy MS, Cunningham S. Bronchiolitis in children: summary of NICE guidance. BMJ: British Medical Journal [Internet]. 2015 [cited 2023 Aug 29];350. Available from: https://www.jstor.org/stable/26520871

14. Fernandes RM, Bialy LM, Vandermeer B, Tjosvold L, Plint AC, Patel H, et al. Glucocorticoids for acute viral bronchiolitis in infants and young children. Cochrane Database Syst Rev. 2013 June 4;2013(6):CD004878.

15. Schober T, Wong K, DeLisle G, Caya C, Brendish NJ, Clark TW, et al. Clinical Outcomes of Rapid Respiratory Virus Testing in Emergency Departments. JAMA internal Medicine. 2024;

16. Pandey M, Lyttle MD, Cathie K, Munro A, Waterfield T, Roland D, et al. Point-of-care testing in Paediatric settings in the UK and Ireland: a cross-sectional study. BMC Emergency Medicine. 2022 Jan 11;22(1):6.

17. BTS_SIGN Guideline for the management of asthma 2019.pdf.

18. Wilson C, Rooshenas L, Paramasivan S, Elliott D, Jepson M, Strong S, et al. Development of a framework to improve the process of recruitment to randomised controlled trials (RCTs): the SEAR (Screened, Eligible, Approached, Randomised) framework. Trials. 2018 Dec;19(1):50.

19. Teresi JA, Yu X, Stewart AL, Hays RD. Guidelines for Designing and Evaluating Feasibility Pilot Studies. Medical Care. 2022 Jan;60(1):95–103.

20. Lee B, Turner S, Borland M, Csonka P, Grigg J, Guilbert TW, et al. Efficacy of oral corticosteroids for acute preschool wheeze: a systematic review and individual participant data meta-analysis of randomised clinical trials. The Lancet Respiratory Medicine [Internet]. 2024 Mar 22 [cited 2024 Mar 25];0(0). Available from: https://www.thelancet.com/journals/lanres/article/PIIS2213-2600(24)00041-9/fulltext#supplementaryMaterial

21. Aschbrenner KA, Kruse G, Gallo JJ, Plano Clark VL. Applying mixed methods to pilot feasibility studies to inform intervention trials. Pilot Feasibility Stud. 2022 Sept 26;8(1):217.

22. Harris PA, Taylor R, Thielke R, Payne J, Gonzalez N, Conde JG. Research Electronic Data Capture (REDCap) - A metadata-driven methodology and workflow process for providing translational research informatics support. J Biomed Inform. 2009 Apr;42(2):377–81.

23. Sekhon M, Cartwright M, Francis JJ. Development of a theory-informed questionnaire to assess the acceptability of healthcare interventions. BMC Health Serv Res. 2022 Dec;22(1):279.

24. Whitehead AL, Julious SA, Cooper CL, Campbell MJ. Estimating the sample size for a pilot randomised trial to minimise the overall trial sample size for the external pilot and main trial for a continuous outcome variable. Stat Methods Med Res. 2016 June;25(3):1057–73.

25. Wilson K, Umana E, McCleary D, Waterfield T, Woolfall K. Exploring communication preferences and risk thresholds of clinicians and parents of febrile infants under 90 days presenting to the emergency department: a qualitative study. Arch Dis Child. 2024 Nov;109(11):886–93.

26. Hewitt-Taylor J. Use of constant comparative analysis in qualitative research. Nursing standard. 2001;15(42):39–42.

27. Moher D, Hopewell S, Schulz KF, Montori V, Gøtzsche PC, Devereaux PJ, et al. CONSORT 2010 Explanation and Elaboration: updated guidelines for reporting parallel group randomised trials. BMJ. 2010 Mar 24;340:c869.

28. Smith CT, Hopkins C, Sydes M, Woolfall K, Clarke M, Murray G, et al. Good practice principles for sharing individual participant data from publicly funded clinical trials. Trials. 2015 Nov 16;16(2):O1.

